# Myeloid-derived suppressor cells in the blood of Iranian COVID-19 patients

**DOI:** 10.1101/2021.07.07.21260141

**Authors:** Esmaeil Mortaz, Mehrnaz Movasaghi, Ali Basiri, Neda K. Dezfuli, Neda Dalil Roofchayee, Hamidreza Jamaati, Johan Garssen, Ian M. Adcock

## Abstract

**Background:** A cytokine storm and lymphopenia are reported in severe acute respiratory syndrome coronavirus 2 (SARS-CoV-2) infection associated with coronavirus disease 2019 (COVID-19). Myeloid-derived suppressive cells (MDSCs) exist in two different forms, granulocyte (G-MDSCs) and monocytic (M-MDSCs) that both suppress T-cell function. Serum IL-6 and IL-8 levels seem to correlate with the number of blood MDSCs.

**Objective:** To determine the frequency of MDSCs in severe COVID-19 patients from Iran and their correlations with serum IL-8 levels.

**Methods:** 37 severe (8 on ventilation, 29 without ventilation) and 13 moderate COVID-19 patients together with 8 healthy subjects were enrolled at the Masih Daneshvari Hospital, Tehran-Iran between 10th April 2020-9th March 2021. Clinical and biochemical features, serum and whole blood were obtained. CD14, CD15, CD11b and HLA-DR expression on MDSCs was measured by flow cytometry.

**Results:** M-MDSCs (P≤0.0001) and G-MDSCs (P≤0.0001) frequency were higher in Iranian COVID-19 patients compared to healthy subjects. M-MDSC frequency was higher in non-ventilated compared to moderate COVID-19 subjects (P=0.004). Serum IL-8 levels were higher in patients with COVID-19 than in normal healthy subjects (P=0.03). IL8 level was significant difference in ventilated, non-ventilated and moderate patients (P=0.005). The frequency of G-MDSCs correlated negatively with INR (r=-0.39, P=0.02).

**Conclusion:** Serum IL-8 levels did not correlate with the number of systemic MDSCs in COVID-19 patients. The highest levels of M-MDSCs were seen in the blood of severe non-ventilated patients. MDSC frequency in blood in the current study did not predict the survival and severity of COVID-19 patients.

## Introduction

The rapid spread of severe acute respiratory syndrome coronavirus 2 (SARS-CoV-2), infection since its recognition in Wuhan China in December 2019 has resulted in a pandemic disease (1). Clinical observations suggest that SARS-CoV-2 infection can range from an asymptomatic infection through a respiratory illness with a fever and a dry cough to a severe acute respiratory distress syndrome-like disease requiring ventilation. There is a high rate of human-to-human transmission (2). In addition, the mortality rate in patients with severe coronavirus disease 2019 (COVID-19) is high (3.4% mortality rate) (3). One of the most challenging complications of severe SARS-CoV-2 infection is a respiratory pneumonia. The pathophysiology of severe diseases appears variable (4).

It is important to find markers that can predict the prognosis of COVID-19 disease. Immunological studies on two other coronavirus infections, SARS-COV1 and MERS-COV have shown that immune-based pathological mechanisms are responsible for disease severity. These pathological mechanisms include lymphopenia and unregulated immune responses (3, 5). Understanding the mechanism(s) and immunological patterns observed in COVID-19 is necessary for the development of effective therapeutic interventions. Myeloid derived suppressive cells (MDSCs) represent an intrinsic portion of the myeloid-cell lineage and are a heterogeneous group of relatively immature myeloid cells (6).

Several studies have described mechanisms of MDSCs mediated immune suppression (6). MDSCs are divided into granulocytic (G-MDSCs or PMN-MDSC) and monocytic (M-MDSCs). In healthy individuals, immature myeloid cells (IMCs) produced in the bone marrow quickly differentiate into mature granulocytes, macrophages or dendritic cells (DCs) [7]. In cancer, sepsis, trauma, bone marrow transplantation and some autoimmune disorders, a partial block in the differentiation of IMCs into mature myeloid cells leads to an expansion MDSCs. MDSC-mediated suppression of certain T-cell function results in an excessive immune response (6, 7). G-MDSCs produce high levels of ROS and low levels of NO whilst the M-MDSC subset produce low levels of ROS and high levels of NO and both subsets produce arginase (8). MDSCs modulated cytokine production (9) and can promote cancer metastasis in humans and in animal models (8, 10).

However, MDSCs may play a beneficial role in acute inflammatory processes such as in dengue virus infection where they decrease the inflammatory response and the subsequent immune-mediated pathology (11). Studies have reported that G-MDSC and M-MDSC expansion occurs in the systemic circulation of mainly patients with severe COVID-19 (3, 12) but these changes require validation different cohort’s of different ethnicity and genetic background. We hypothesised, that the frequency of MDSCs and the potential to induce immune suppression is related to the severity of COVID-19 disease in Iranian subjects. Thus, we studied the frequency of M-MDSCs and G-MDSCs in the systemic circulation of in moderate and severe Iranian COVID-19 patients and determined whether this was associated with systemic inflammation as reflected by IL-8 levels.

## Materials and Methods

### Patients

Fifty confirmed COVID-19 patients including 8 severe patients on ventilation, 29 severe non-ventilated and 13 moderate patients were enrolled into the study upon admission to the Masih Daneshvari Hospital of Shahid Beheshti Medical University (Tehran-Iran) between 10th April 2020-9th March, 2021. All patients were diagnosed according to World Health Organization interim guidance (13). Severe COVID-19 disease was confirmed by the presence of at least one of the following: respiratory rate≥30/min; blood oxygen saturation ≤93%; ratio of partial pressure of oxygen in arterial blood to the inspired oxygen fraction (PaO2FiO2) <300; lung infiltrates present on>50% of the lung field (14). Eight healthy age-matched controls were also recruited. This study was approved by the institutional ethics board of Masih Daneshvari Hospital ethical committee <u>(IR.SBMU.NRITLD.REC.1399.122</u>).

### Data collection

The clinical records of patients were interpreted by the research team of the Department of Critical Care Medicine, Masih Daneshvari Hospital of Shahid Beheshti University. Clinical, laboratory, and radiological properties and treatments and outcomes data were collected from electronic medical records. The information recorded included demographic data, medical history, underlying comorbidities, symptoms, signs, laboratory findings; chest computed tomographic (CT) scans, and treatment measures including antiviral therapy, corticosteroid therapy, respiratory support and kidney replacement therapy.

### Laboratory examination of blood samples

Whole blood samples containing anti-coagulant EDTA (3ml) and with citrate (3ml) or no anticoagulant (3ml) were obtained from all participants upon admission. Tubes containing blood without anticoagulant were centrifuged, the serum collected and stored at -80°C for IL-8 measurement. Serum biochemical tests including kidney and liver function tests: creatinine kinase-muscle/brain activity (CK-MB), lactate dehydrogenase (LDH), C-reactive protein (CRP), ferritin, creatine phosphokinase (CPK), and the international normalized ratio (INR) were also performed. The erythrocyte sedimentation ratio was determined in citrate-treated whole blood samples.

### Cell staining and flow cytometry

Whole blood samples (3 ml) were collected in EDTA-treated tubes and MDSCs were analysed by flow cytometry (FACS Calibour, BD, USA). Whole blood samples were then stained with anti-CD11b APC (BD Biosciences, CA, USA), anti-HLA-DR PE (eBioscience, San Diego, CA, USA), anti-CD14 PerCP-cy5.5 (eBioscience) and anti-CD15 FITC (BD Biosciences) for 30 min in the dark as described before (15, 16). Cells were then washed and suspended in FACS buffer before 20,000 events were analysed by FACS. The gating strategy was CD11b+/HLA-DR-/dim and within this population CD14+/CD15-cells and CD14-/CD15+ were identified as described previously (15, 16). Flow cytometry analysis was calculated as the frequency (percentage) of the respective subset of leukocytes and the data was subsequently analysed by FlowJO-V10 software (USA).

### Measurement of IL-8

Serum concentrations of IL-8 were measured by linked immunosorbent assay (ELISA) (BD Biosciences, CA, USA) according to the manufacturer’s protocol.

### Statistical analysis

Analysis of data was performed using the SPSS program version 16.0 (SPSS, Inc. Chicago, USA) and Graph Pad Prism software (version 6; 07 Graph Pad Software, Inc.). Results were presented as the mean ± standard deviation (SD). The measurement data between two groups were analyzed using Student’s *t*-test. The non-parametric Mann-Whitney U test was used for non-normally distributed variables. Difference among multiple groups was compared using one⍰ way analysis of variance (ANOVA), p<0.05 was considered as statistically significant.

## Results

### Demographic and clinical characteristics of patients with COVID-19 and healthy control subjects

The demographic and clinical characteristics of the participants are shown in **Table 1**. There were no significant differences in the serum levels of ESR, CRP, LDH, Troponin, and MB activity between ICU (ventilated and non-ventilated) and non-ICU moderate COVID-19 patients. However, CPK was higher in ICU patients (ventilated and non-ventilated) than non-ICU moderate patients (P=0.01). (**Table 1**).

**Table 1:**
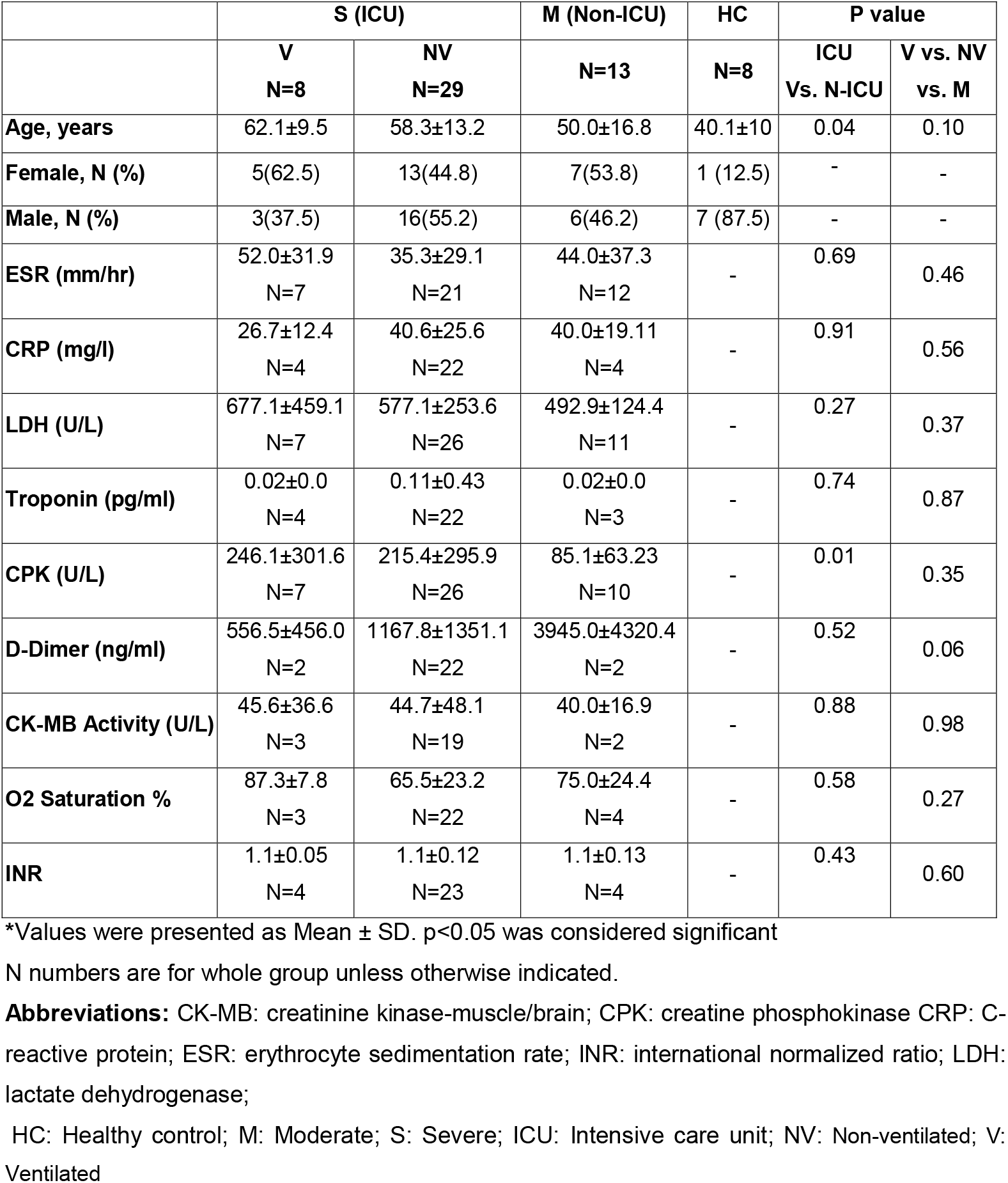
The demographic data and biochemical characters of participants.

### MDSC analysis

The frequency of M-MDSCs (HLA-DR^-/dim^CD11b^+^CD14^+^CD15^-^ cells) and G-MDSCs (HLA-DR^-/dim^CD11b^+^CD14^-^CD15^+^ cells) in the blood were evaluated as described previously (15). The gating strategy adopted and the frequency of G-MDSC and M-MDSC cells in representative healthy control subjects and COVID-19 patients of differing severity are shown in **Fig. 1**. There was a significantly greater frequency of M-DSCs (**Fig. 2A, Table 2**) and G-MDSCs (**Fig. 2B, Table 2**) in all COVID-19 patients as a group compared to HC (P≤0.0001, P≤0.0001, respectively).

**Figure 1:**
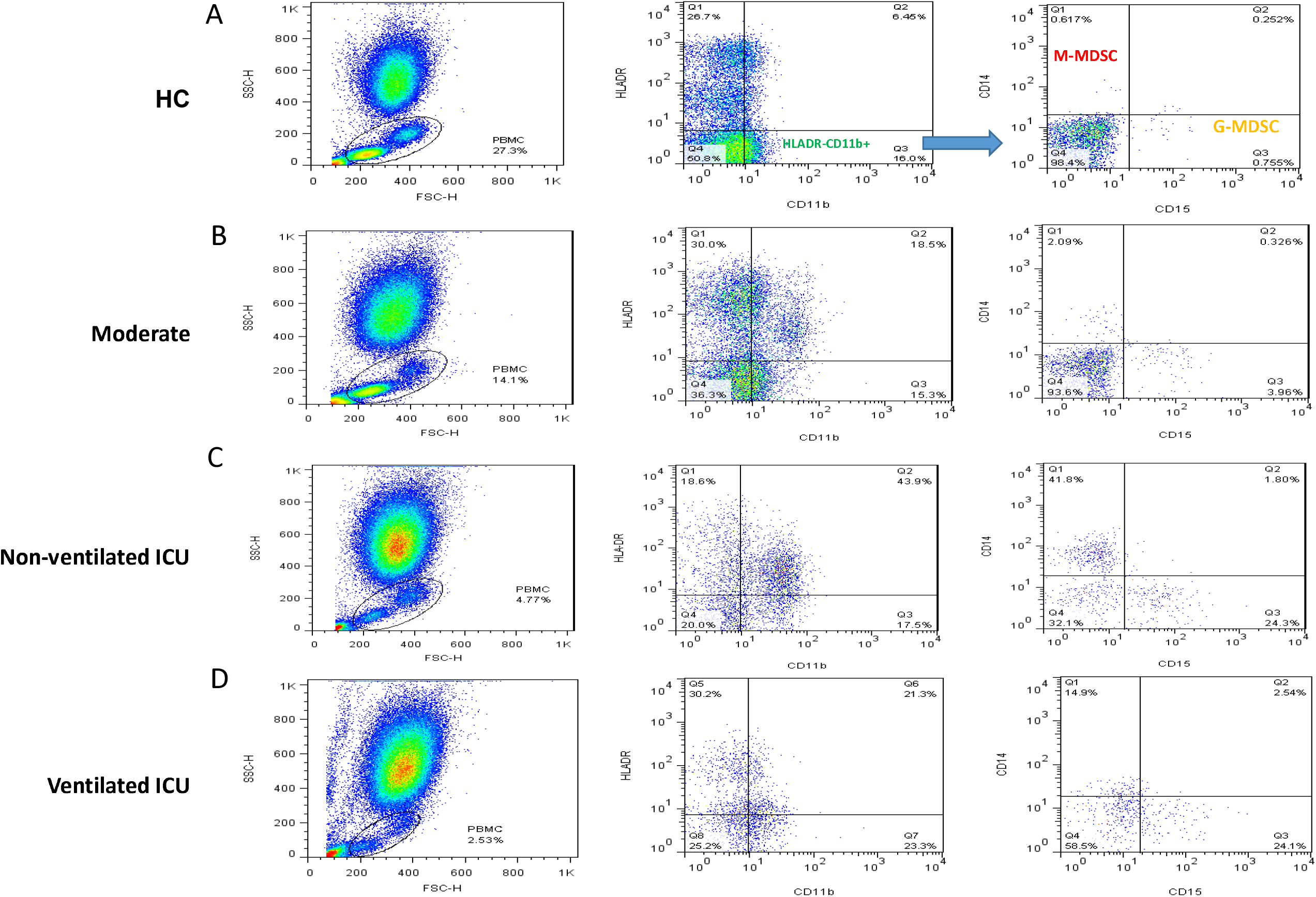
Gating strategy for M-MDSC and G-MDSC cells. Representative dot plots of flow cytometry data M-MDSCs (HLA-DR^-/dim^CD11b^+^CD14^+^CD15^-^ cells) and G-MDSCs (HLA-DR^-/dim^CD11b^+^CD14^-^CD15^+^ cells) in the blood of a HC (A) and moderate (B) non-ventilated ICU (C) and ventilated ICU (D) patients frequency of cells are indicated in the text boxes in the top left (M-MDSC) and bottom right (G-MDSC) panels.

**Figure 2:**
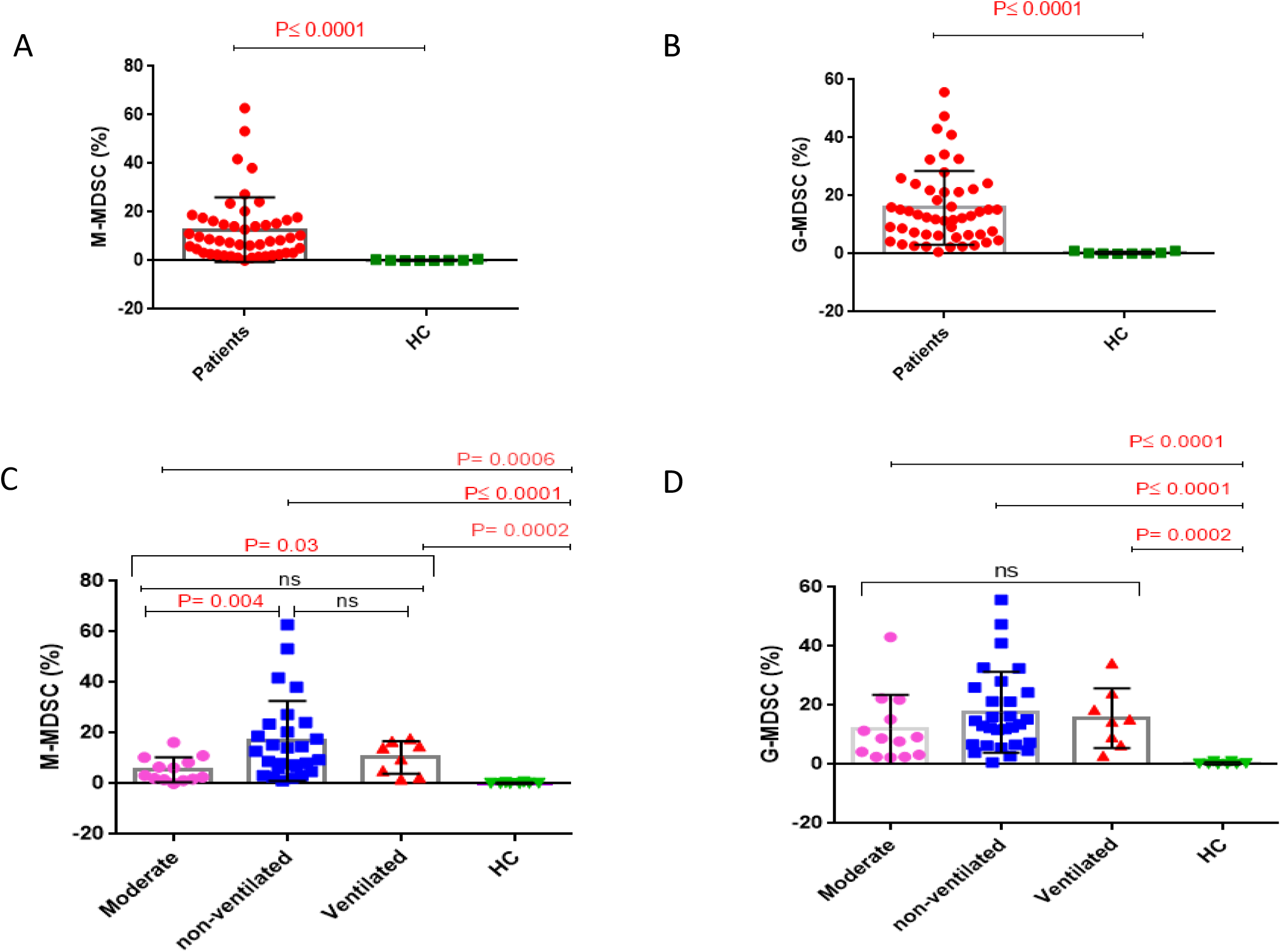
Frequency of M-MDSCs and G-MDSCs in COVID-19 patients and healthy controls subjects. Dot plot of the frequency of M-MDSCs (A) and G-MDSCs (B) between COVID-19 patients compared to HC. C) Frequency of M-MDSCs individually was calculated. Individual values for each subject with the mean (SD) in severe COVID-19 patients admitted to the intensive care unit (ICU) who were either ventilated or non-ventilated or who had moderate disease and were not admitted to the ICU presented as dot plot. (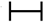: shows significant between two groups by using t-test or Mann-Whitney U test, ⎴: shows statistic differences among ventilated, non-ventilated and moderate patients by using one⍰ way analysis of variance) D) Frequency of G-MDSCs individually was calculated and presented as dot plot. (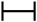: shows significant between two marked groups groups. ⎴ ; shows significant between two groups by using t-test or Mann-Whitney U test ⎴ : shows statistic differences among ventilated, non-ventilated and moderate patients by using one⍰ way analysis of variance

**Table 2:**
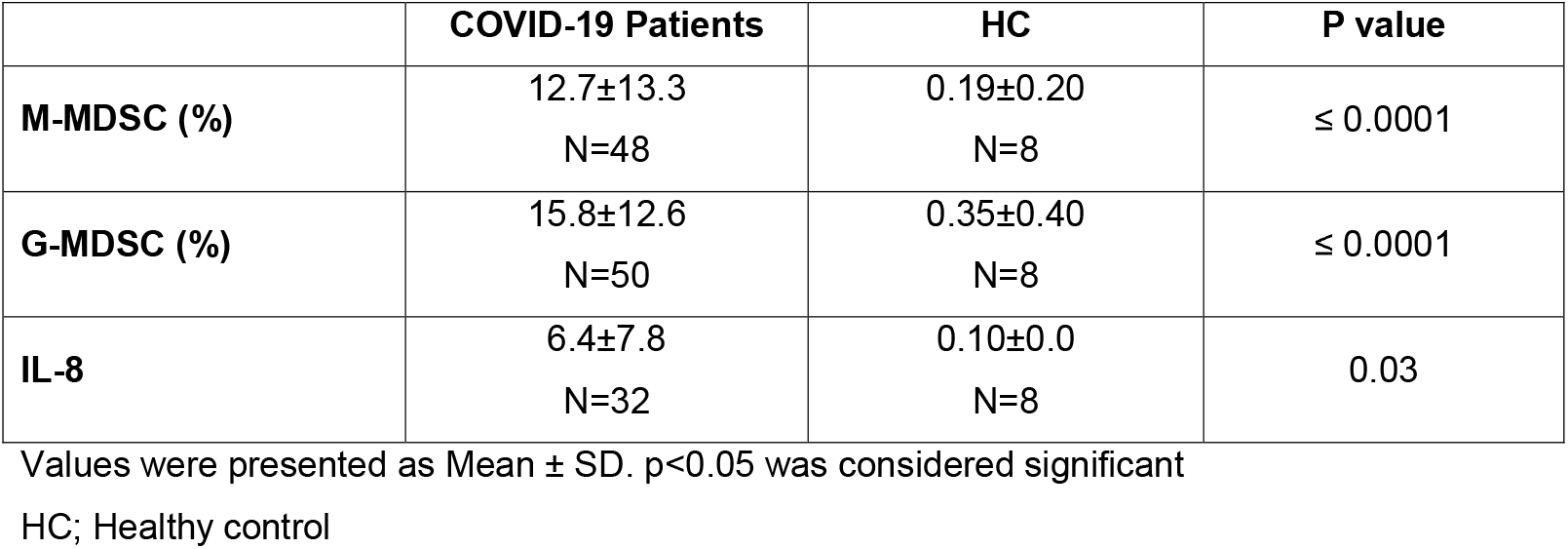
The frequency of MDSCs and serum levels of IL-8 in COVID-19 patients and healthy controls

|  | **COVID-19 Patients** | **HC** | **P value** |
| --- | --- | --- | --- |
| **M-MDSC (%)** | 12.7±13.3<br>N=48 | 0.19±0.20<br>N=8 | ≤ 0.0001 |
| **G-MDSC (%)** | 15.8±12.6<br>N=50 | 0.35±0.40<br>N=8 | ≤ 0.0001 |
| **IL-8** | 6.4±7.8<br>N=32 | 0.10±0.0<br>N=8 | 0.03 |

Values were presented as Mean ± SD. p<0.05 was considered significant
HC; Healthy control

Next we analysed the M-MDSC and G-MDSC cells in moderate and severe COVID-19 patients. There are significant differences in the frequency of M-MDSCs between moderate, non-ventilate and ventilated patients (P=0.03)(**Fig. 2C, Table 3**). In addition, there was a significant increase in M-MDSC frequency in severe COVID-19 patients who were not ventilated compared to moderate patients (P=0.004) (**Fig. 2C, Table 3**). In contrast, no significant differences were found between ventilated and non-ventilated patients in the frequency of M-MDSC (**Fig. 2C**). Furthermore, moderate and severe COVID-19 patients have a significantly higher frequency of M-MDSC compared with HC (**Fig.2C, Table 3**).

**Table 3:** The percentage of MDSCs and levels of serum IL-8 in severe COVID-19 patients according to ICU status

|  | HC | M | S |  |  | P value |  |  |  |  |  |
| --- | --- | --- | --- | --- | --- | --- | --- | --- | --- | --- | --- |
|  |  |  | NV | V | M vs.<br>HC | NV vs.<br>HC | V vs.<br>HC | V vs. NV<br>vs. M | NV vs.<br>M | V vs. M | NV vs.<br>V |
| M-MDSC (%) | 0.19±0.20 | 5.4±4.8<br>N=13 | 16.8±15.8<br>N=27 | 10.3±6.4<br>N=8 | 0.0006 | ≤0.0001 | 0.0002 | 0.03 | 0.004 | 0.06 | 0.43 |
| G-MDSC (%) | 0.35±0.40 | 11.8±11.6<br>N=13 | 17.6±13.6<br>N=29 | 15.6±10.0<br>N=8 | ≤0.0001 | ≤0.0001 | 0.0002 | 0.40 | 0.19 | 0.46 | 0.70 |
| IL-8 | 0.10±0.00 | 2.8±3.0<br>N=10 | 5.7±2.7<br>N=17 | 15.7±16.7<br>N=5 | 0.02 | ≤0.0001 | 0.02 | 0.005 | 0.01 | 0.008 | 0.04 |

Values were presented as Mean ± SD. p<0.05 was considered significant.
**Abbreviations:**
HC: Healthy Controls
M: Moderate
S: Severe
V: Ventilated
NV: Non Ventilated

No significant differences in G-MDSC cell frequencies were found between moderate and severe COVID-19 patients although all COVID-19 groups had a higher frequency than observed for HC (**Fig. 2D, Table 3**).

### IL-8 levels

The serum levels of IL-8 in COVID-19 patients (moderate and severe) was significantly higher than in healthy control subjects (P=0.03) (**Fig. 3A, Table 2**). In addition, serum IL-8 levels were significantly higher in severe COVID-19 patients on ventilation compared to those who were not ventilated (P=0.04) and in moderate patients (P=0.008) (**Fig. 3B, Table 3**). Systemic IL-8 levels were higher in non-ventilated patients than in patients with moderate COVID-19 (P= 0.01) (**Fig. 3B, Table 3**).

**Figure 3:**
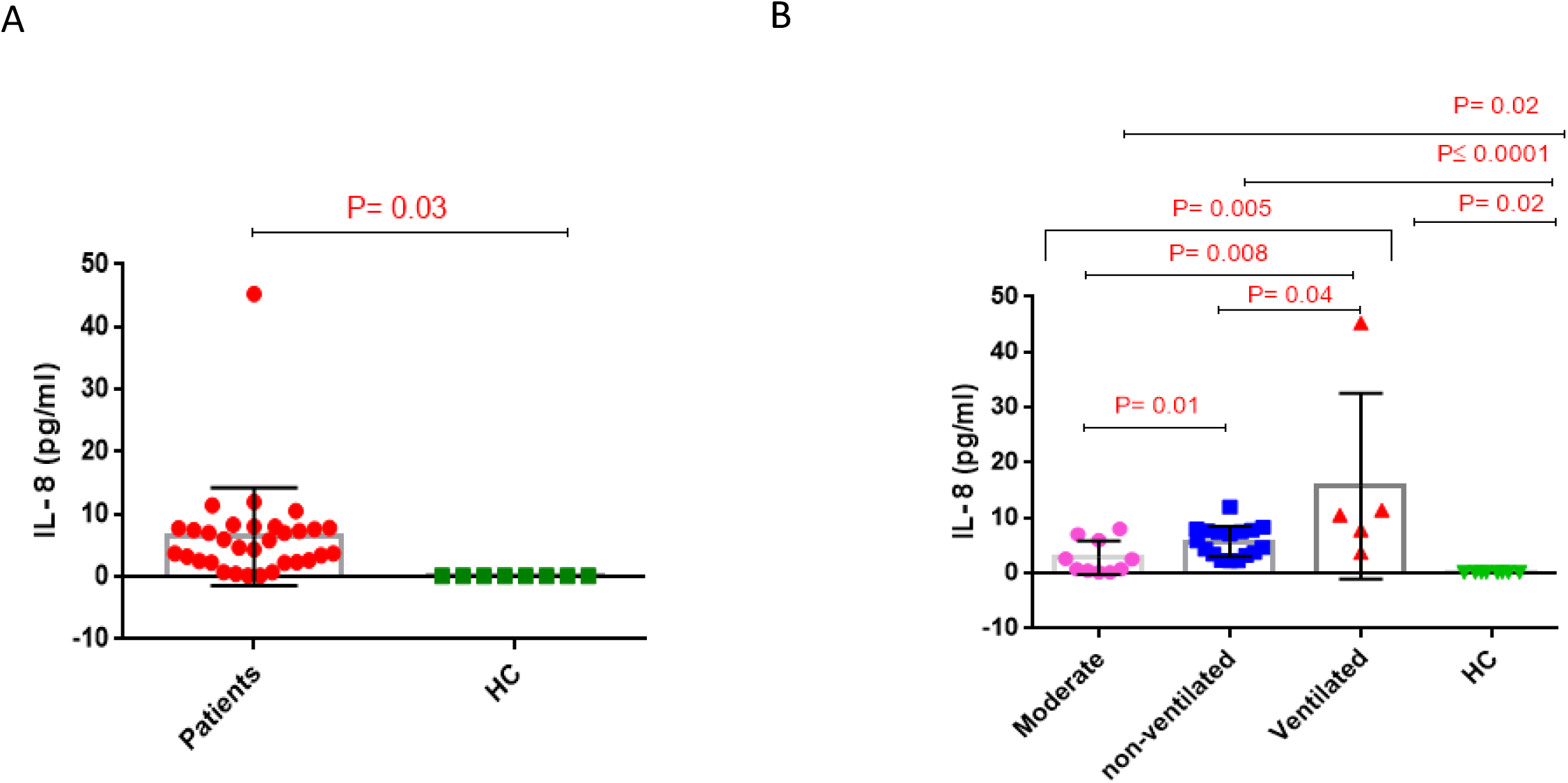
IL-8 level in COVID-19 patients. Serum IL-8 concentration in COVID-19 patients versus HC (A). Serum IL-8 levels in severe COVID-19 patients admitted to the intensive care unit (ICU) who were either ventilated or non-ventilated or had moderate disease (B). Results are presented as dot plots of individual values for each subject with the mean (SD) (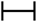 : shows significant between two groups by using t-test or Mann-Whitney U test, ⎴ : shows statistic differences among ventilated, non-ventilated and moderate patients by using one⍰ way analysis of variance).

### Correlation between M-MDSCs/G-MDSCs/IL-8 with clinical and serum parameters outcome

The relationships between M-MDSCs and G-MDSCs frequency and IL-8 concentration with serum biochemical markers LDH, ESR, CRP, troponin, CPK, CK-MB activity and D-dimer, INR, age and arterial O2 saturation were evaluated. G-MDSC frequency correlated negatively with INR (r=-0.39, P=0.02) and no other significant correlations were observed (**Table 4**).

**Table 4:**
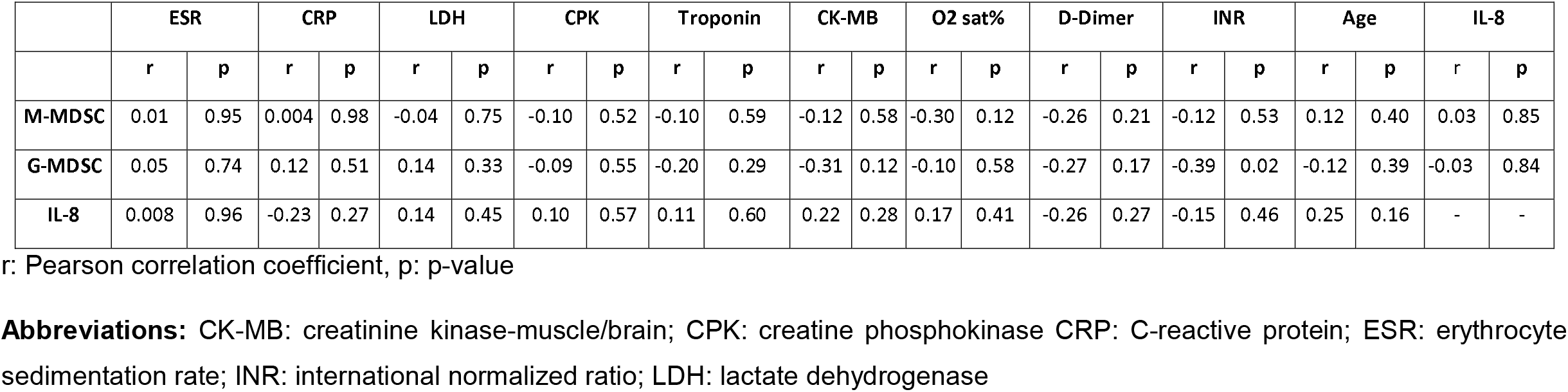
Correlation analysis between M-MDSC, G-MDSC, IL-8 and other factors in COVID-19 patients.

|  | ESR |  | CRP |  | LDH |  | CPK |  | Troponin |  | CK-MB |  | O2 sat% |  | D-Dimer |  | INR |  | Age |  | IL-8 |  |
| --- | --- | --- | --- | --- | --- | --- | --- | --- | --- | --- | --- | --- | --- | --- | --- | --- | --- | --- | --- | --- | --- | --- |
|  | r | p | r | p | r | p | r | p | r | p | r | p | r | p | r | p | r | p | r | p | r | p |
| **M-MDSC** | 0.01 | 0.95 | 0.004 | 0.98 | -0.04 | 0.75 | -0.10 | 0.52 | -0.10 | 0.59 | -0.12 | 0.58 | -0.30 | 0.12 | -0.26 | 0.21 | -0.12 | 0.53 | 0.12 | 0.40 | 0.03 | 0.85 |
| **G-MDSC** | 0.05 | 0.74 | 0.12 | 0.51 | 0.14 | 0.33 | -0.09 | 0.55 | -0.20 | 0.29 | -0.31 | 0.12 | -0.10 | 0.58 | -0.27 | 0.17 | -0.39 | 0.02 | -0.12 | 0.39 | -0.03 | 0.84 |
| **IL-8** | 0.008 | 0.96 | -0.23 | 0.27 | 0.14 | 0.45 | 0.10 | 0.57 | 0.11 | 0.60 | 0.22 | 0.28 | 0.17 | 0.41 | -0.26 | 0.27 | -0.15 | 0.46 | 0.25 | 0.16 | - | - |

r: Pearson correlation coefficient, p: p-value
**Abbreviations:** CK-MB: creatinine kinase-muscle/brain; CPK: creatine phosphokinase CRP: C-reactive protein; ESR: erythrocyte sedimentation rate; INR: international normalized ratio; LDH: lactate dehydrogenase

## Discussion

We demonstrated that M-MDSCs and G-MDSCs were higher in the peripheral blood of COVID-19 patients from Iran compared to healthy controls. There was a significant increase in the presence of M-MDSCs in non-ventilated compared to moderate COVID-19 patients. Overall, serum IL-8 levels were significantly higher in COVID-19 patients compared to healthy controls and in severe ventilated COVID-19 patients compared to non-ventilated COVID-9 patients.

MDSCs are immature myeloid cells that suppress T cell responses (8). M-MDSC and G-MDSCs expressed the IL-8 receptors CXCR1 and CXCR2 and, therefore, IL-8 may attract peripheral MDSCs to tumour sites (17). In addition, increased levels of IL-8 have been associated with more severe COVID-19 (18) although one study has reported that blood levels of IL-8 and MDSCs in ICU and non-ICU COVID-19 patients were similar (19). Bourboulis and colleagues showed that COVID-19 patients with severe respiratory failure presented with a lower percentage of HLA-DR on CD14 monocytes (20). Furthermore CD14+ HLA-DR^low^ cells were seen in severe COVID-19 patients and associated immune dysregulation (20). This MDSC expansion seen may account for the T cells dysregulation, particularly that of CD8 T cells, seen in COVID-19 patients who develop acute respiratory distress syndrome (21).

Agrati and co-workers showed that the percentage of G-MDSCs was increased in severe and mild COVID-19 patients (18). In contrast, COVID-19 patients during the later convalescent phase of severe disease showed a decreased frequency of MDSC cells which was linked to higher systemic levels of IL-8, IL-1β and TNF-α and reduced TGF-β levels (18). Systemic IL-8 levels positively correlated with the duration of COVID-19 and is an important neutrophil chemoattractant with neutrophilia and the neutrophils/lymphocyte ratio (NLR) being markers of SARS-COV-2 infection (22).

We have demonstrated increased numbers of G-MDSC cells in moderate and severe COVID-19 patients and healthy control subjects but we did not find any significant difference between moderate and severe patients. In a study from Italy, PMN-MDSC frequency was higher in severe COVID-19 patients who died upon admission to hospital compared to those who survived (23). This study also reported that plasma IL-8 levels were also higher in non-survivors than in survivors upon admission, and decreased to levels comparable to those of survivors at later time points (23). However, the higher levels of serum IL-8 in ventilated COVID-19 patients in our study are consistent with other reports (23). We are unable to account for the discrepancy in the frequency of G-MDSC cells in ventilated severe COVID-19 patients from Iran compared to Italy but suggest that this may reflect differences in treatment and/or ethnicity between subjects in Iran and Italy or possibly the number of subjects studied. Overall, our data does not support the hypothesis that MDSCs are a predictor of COVID-19 outcomes.

In the current study we have also seen a difference in IL-8 levels between ventilated and non-ventilated severe COVID-19 patients and that both groups had higher IL-8 levels than in moderate disease and in healthy controls. Furthermore, there were no differences in peripheral blood G-MDSC frequency between COVID-19 patients with different severities although they were all groups of patients were higher than healthy control subjects. In contrast, the frequency of M-MDSC in the blood of non-ventilated severe COVID-19 patients was higher than patients with moderate disease.

We did not find any link between systemic IL-8 levels and the proportion of MDSC cells in blood. This suggests that either other factors are responsible for the production of MDSCs in these patients or that the enhanced blood levels of IL-8 reflect much higher levels in the lung and other COVID-19-infected tissues and this results in margination to these tissues. This may be even more evident in patients with very severe disease who are on ventilation where margination of M-MDSCs from the blood to the lung or kidney results in suppression of the local immune response. Future studies are required using matched peripheral blood and lung samples, for example, to determine whether this is indeed the case. A recent study indicated that M-MDSC frequency is enhanced in blood but not in the upper airways of COVID-19 patients (12).

In HIV-1 patients, the expansion of MDSCs promotes the differentiation of regulatory T cells and thereby control T cell differentiation and function (24). Thus, in HIV-1 patients MDSCs play a role in countering viral persistence and may suggest a new immunotherapeutic strategy against human viral diseases (24). Moreover, increased levels of M-MDSCs strongly correlates with a worse outcome in septic shock (25). MDSCs play a dual role in patients with sepsis (26) being beneficial by reducing inflammation during early stage disease stage whilst in the latter stages of disease they are able to induce long term immunosuppression (18, 26).

In summary, our data in Iranian patients are in line with other studies showing increased IL-8 in the peripheral blood of COVID-19 patients and that higher expression was associated with worse clinical outcomes. This may lead to the expansion and recruitment of MDSC cells and that targeting this cytokine-MDSC network may provide a new approach to improve clinical outcomes of COVID-19 patients.

## Data Availability

NO applicable

## Declarations and footnotes

### ETHICS STATEMENT

The studies involving human participants were reviewed and approved by Masih Daneshvari Hospital ethical Board. The patients/participants provided their written informed consent to participate in this study.

### AUTHOR CONTRIBUTIONS

EM, NKD and NDR did experiments. EM wrote the draft of paper. MM, AB, HG, JG revised the paper. IA approved the final version as corresponding author. All authors contributed to the article and approved the submitted version.

## ACKNOWLEDGMENTS

We acknowledge all study participants whom are alive and remember those patients in this study who died of COVID-19.

